# Diverging Pre-Pandemic Mortality Trends: Age-Specific and Cause-Specific Patterns Across High-Income Countries

**DOI:** 10.64898/2026.06.01.26354619

**Authors:** Francisco J. Pérez-Reche, Jennifer Summers, Gareth T. Jones, Gary J. Macfarlane

**Affiliations:** School of Natural and Computing Sciences, University of Aberdeen, Old Aberdeen, AB24 3UE, Aberdeen, United Kingdom; Epidemiology Group, School of Medicine, Medical Sciences and Nutrition, University of Aberdeen, Aberdeen, United Kingdom

**Keywords:** Mortality rates, Age-specific mortality, Cause-specific mortality, External causes of death, Ill-defined causes of death, International comparisons

## Abstract

**Background:** Mortality improvement has slowed or reversed in several high-income countries, but cross-national evidence on the causes of death underlying these trends is limited.

**Methods:** We analysed all-cause and cause-specific mortality in 32 high-income countries during 2012–2019, using causes as certified and coded to ICD-10 chapters and three-character codes. Annual percentage changes in mortality rates were estimated by Poisson regression accounting for overdispersion and adjusting for age composition. Changes were classified as material if ≥1% per year and survived false discovery rate control, as stagnation if significant but smaller or non-significant with a confidence interval within ±1%, and as inconclusive otherwise.

**Results:** Material all-cause mortality increases were confined to the United States and Canada at ages 15–44, and France in infancy. None occurred from age 45 onwards in any country. Stagnation was most extensive in the United Kingdom. Cause-specific increases were more widespread: most countries had at least one rising ICD-10 chapter. Patterns varied by age: external causes at younger ages, alcoholic liver disease and hypertensive heart disease at 45–74, and dementia and neurodegenerative disease at the oldest ages, rising in up to 19 countries. Ill-defined causes (R99) rose in 17 countries at 75 and over. At that age, all-cause mortality fell materially in 24 countries, yet 22 still had at least one rising ICD-10 chapter.

**Conclusion:** Adverse cause-specific trends extended beyond the countries and ages where all-cause mortality rose, meaning all-cause surveillance alone would not detect them. Rising ill-defined causes leave part of this change poorly characterised.

## 1. Introduction

Mortality rates declined substantially in most high-income countries through the twentieth century and into the twenty-first^1–4^. However, recent evidence points to increasing divergence in mortality trajectories across these settings^5,6^, challenging the assumption of uniformly improving health outcomes. Countries such as Japan, Switzerland, and the Republic of Korea have continued to experience broad improvements into the 2010s^7,8^, whereas others, notably the United States (US), the United Kingdom (UK), and Canada, have seen a slowdown in the decreases or a shift from declining to increasing mortality, particularly at working ages^9–13^. The extent to which these patterns are shared across countries, and the specific causes underlying them, remains incompletely understood.

Recent analyses highlight a concerning pattern in the UK, where all-cause mortality among adults aged 30–54 years has increased by up to 1.4% annually since around 2012 ^13^. Within the UK, this pattern is especially pronounced in Scotland, and the increases are disproportionately concentrated in socioeconomically deprived communities, contributing to widening health inequalities across the UK^14–17^.

These patterns likely reflect socioeconomic, behavioural and healthcare-related factors. Austerity has the strongest cross-national evidence base^18–20^, with panel analyses linking reductions in government and social spending to adverse mortality trends, though a systematic review rated its certainty as low^21^; austerity acts partly through income inequality and pressure on health and social care provision, which are hard to separate from it^22^.

Influenza and obesity have been quantified and explain a minority of the stall^23–25^. Cardiovascular improvement has stalled in most affected countries, with rising external- cause mortality at young and mid-life ages in the worst performers^5^.

Cross-national evidence is particularly limited for causes of death as certified, rather than the exposures or social and economic conditions that may underlie them. Studies comparing many countries have largely focused on all-cause mortality or life expectancy^5,19,26,27^. Cause- specific analyses, by contrast, have often examined only one or two countries^11,23,28^. Other studies have combined broad international coverage with cause-specific data, but have either grouped causes in advance^6^ or used modelled estimates in which ill-defined causes are redistributed^29,30^. These approaches prevent the study of such causes in their own right.

This study examines age-specific mortality trends across 32 high-income countries between 2012 and 2019. We analyse all-cause and cause-specific mortality at both ICD-10 chapter and individual-code levels, using causes as certified. Although these patterns cannot identify the determinants of mortality trends, they constrain possible explanations: any account of a trend must ultimately explain the changes observed in causes of death.

## 2. Methods

### 2.1 Data sources

All-cause mortality and population exposure data were obtained from the Human Mortality Database^31^ (HMD) for the period 2012–2019. The study period was restricted to pre- pandemic years to avoid the influence of COVID-19 on mortality trends, including potential disruptions to cause-of-death coding practices that could obscure trends in non-COVID causes. Cause-specific mortality data were obtained from the World Health Organization (WHO) Mortality Database^32^ and coded according to the International Classification of Diseases, Tenth Revision (ICD-10)^33^. Countries were eligible if they were represented in both the HMD and WHO databases, and if all eight years from 2012 to 2019 were available in each source. Thirty-two countries met these conditions. All are currently high-income countries (Supplementary File 1, Section 1).

### 2.2 Age groups and cause-of-death classification

Mortality rates were analysed for seven age groups: under 1 year, 1-14, 15–29, 30–44, 45– 59, 60–74, and 75+ years. Infancy is reported separately from childhood because the two have largely distinct causes of death. Cause-specific mortality was examined at both the ICD-10 chapter level and the individual ICD-10 code level. Throughout, ill-defined causes refers to ICD-10 Chapter XVIII (R00–R99), in particular R99, which records that the underlying cause could not be determined rather than a disease process.

### 2.3 Trend estimation

Mortality trends were modelled using Poisson regression with a log link and person-years of exposure as an offset, allowing for overdispersion in annual death counts^34,35^. The model adjusted for changes in age composition within each broad age group. Annual percentage changes (APCs) in mortality rates were estimated from the fitted log-linear trends, with uncertainty quantified using quasi-Poisson standard errors and 95% confidence intervals.

P-values were adjusted for multiple comparisons using the Benjamini–Hochberg procedure (adjusted value *q*), controlling the false discovery rate at 5% within each family of tests^36^. Trends were classified as materially increasing or decreasing only when *q* < 0.05 and |APC| ≥ 1% per year. Remaining cells were classified as significant but small (q < 0.05, absolute APC below 1% per year), as showing no material change (q ≥ 0.05, confidence interval within ±1% per year), or as inconclusive (q ≥ 0.05, interval admitting a larger change). The first two are reported as stagnation; the third is absence of evidence rather than evidence of stability. The 1% per year threshold is on the order of the annual improvement typical of these populations.

Further methodological details are provided in Supplementary File 1.

## 3. Results

### 3.1 Divergence in All-Cause Age-Specific Mortality Trends

Age-specific all-cause mortality trends highlight a critical divergence between nations maintaining health progress and those with increasing mortality rates. Many countries show material declines (≥ 1%/year, surviving control of the false discovery rate) in several age groups. In particular, Japan and the Republic of Korea are the only two to decline materially in every age group (blue cells with a dot in Fig. 1).

**Figure 1.**
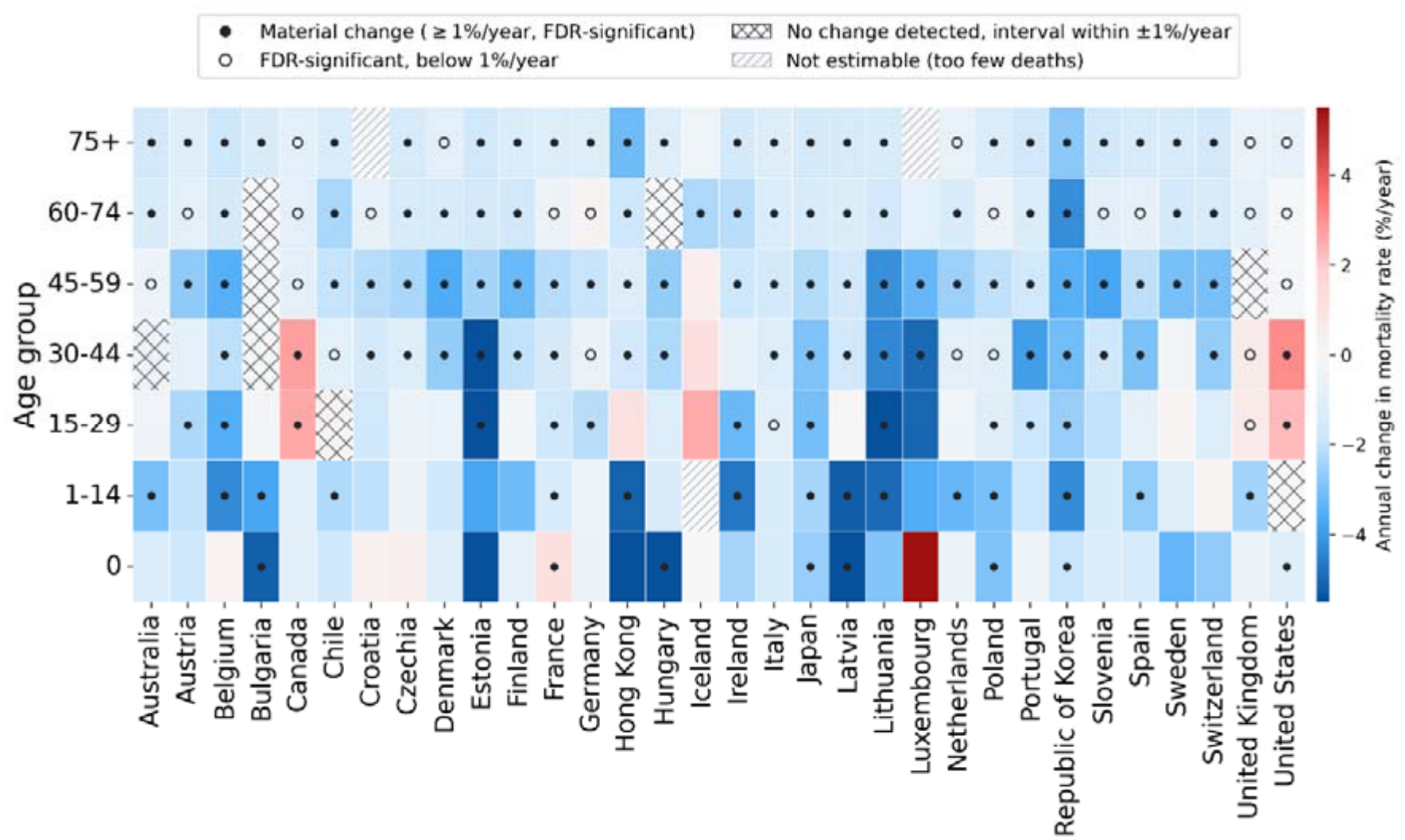
Trends in all-cause mortality rates by age group and country, 2012–2019. Colour shows the annual percentage change in the mortality rate adjusted for age composition within each broad band. A filled dot within a cell marks a material change (FDR-adjusted p-value below 0.05 and |APC| > 1%/year). An open circle marks a change that survives the same control but is estimated below 1% per year. Cross-hatching marks a cell in which the rate confidence interval lies entirely within 1% per year either side of zero, so a change of material size is excluded. A cell with none of these marks is one in which the deaths available could not resolve a trend of that size; it is an absence of evidence, not evidence of stability. Diagonal hatching marks cells that could not be estimated.

Material increases in mortality rates occurred in the US and Canada for the 15-29 and 30–44 age groups, and in France for infants (red cells with a dot in Fig. 1). Increasing mortality rates are also observed for Iceland and the UK for the 15-29 and 30–44 age groups. However, such increases are inconclusive for Iceland and below 1% per year for the UK.

Except for Japan and the Republic of Korea every country has at least one age group in which mortality did not change materially (cells without a filled dot in Figure 1). Stagnation is most extensive in the UK, in five of seven age groups, followed by the US with four and Canada and Bulgaria with three each. Stagnation is of greatest concern where it coincides with high mortality: the US, Bulgaria, Chile and Poland each stagnate in two or more age groups while their standardised rates exceed the age-group median in several of those age groups (Supplementary File 1, Section 3.3). Australia also stagnates in two age groups, but from rates below the median. Trends compared between men and women using a sex-by- year interaction (Supplementary File 1, Section 7) resolved a difference in only 14 of 216 country-by-age cells, all at ages between 30 and 74. In 11 of the 14 cells, mortality fell in both sexes but more slowly in women. The exceptions are the US and Canada at 30–44, where it rose in both and faster in men (by 1.25 and 2.05 percentage points per year), and Iceland at 60–74, where women’s rate fell unusually fast while men’s was essentially flat, leaving men with an additional 3.96 percentage points per year.

### 3.2 Trends by ICD-10 Chapter

Chapter-level trends are summarised in Figure 2, and given in full in the Chapter counts tab of Supplementary File 2. Inconclusive cells are the majority at every age, falling from 84% of those shown at ages 15–29 to 48% at 75 and over, so each count below is a lower bound on how many causes materially varied in the studied period.

**Figure 2.**
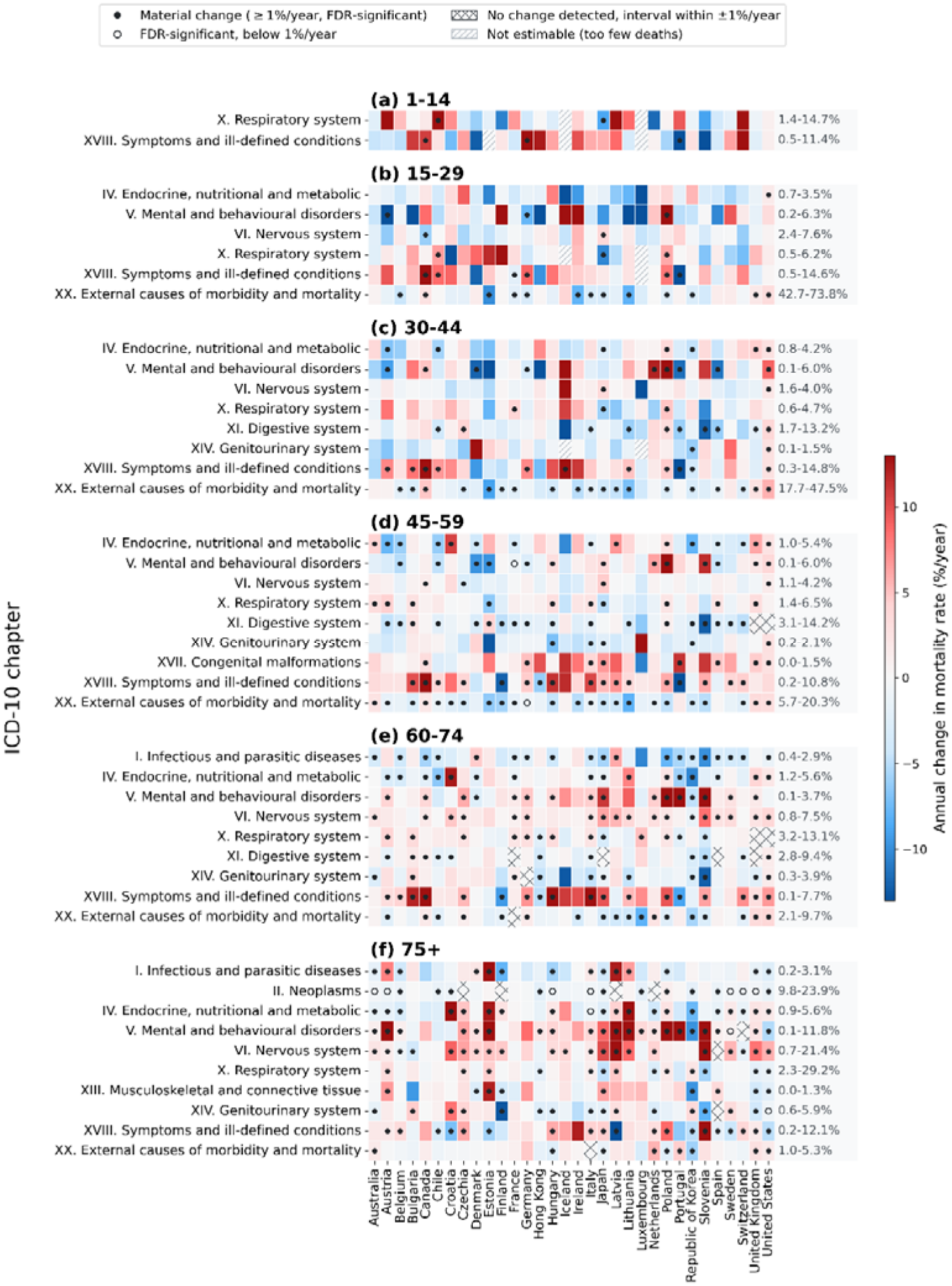
Age-adjusted trends in cause-specific mortality by ICD-10 chapter and country, by age group. Colour, and all four marks, as in Figure 1. Within each age group, a chapter is shown where at least one country had a significant increase, and the chapter accounts for at least 1% of deaths in at least one country. The range at the right of each row gives the smallest and largest share of deaths that chapter reaches across countries. Infants are not shown, as no chapter met both conditions below age 1.

No country had an increasing chapter in infancy, and only three (Canada, Chile and Germany, with one chapter each) at ages 1–14. Across all chapters assessed, an average of 13 of 16 per country are inconclusive at these ages. This is a limit of what cause-of-death data can support when deaths are few, not evidence that childhood mortality patterns were stable.

Increases become resolvable from adolescence onward but remain concentrated. At ages 15–29, seven of 32 countries had at least one increasing chapter, led by Poland with three; in the US, Canada and Chile two chapters rose in each, and in the US and Canada these included external causes, which carry 74% and 69% of deaths respectively at that age. By ages 30–44 fourteen countries had at least one increasing chapter, with the US clearly ahead at seven, followed by Poland with five and Canada with three.

From ages 45-59 onward, rising chapters become common, and the countries carrying them change. At 45–59, 23 of 32 countries had at least one rising chapter, and rising chapters averaged 1.7 per country across all 32. The US had six, Poland and the UK five each. At 60– 74 the average was 2.3 per country, with the US again at six and Czechia, Germany and Canada at five each. At 75 and over it was 2.8 per country, and the four leading countries (Austria, Czechia, Estonia and Latvia with six each) were all in central or northern Europe, the US having fallen to three.

Differences between men and women were resolved in 56 of the 2,834 estimable chapter- by-country cells, none below age 45. Neoplasms and respiratory disease account for 33 of them, and in most cases women’s mortality fell more slowly than men’s.

### 3.3 Trends by ICD-10 Code

Because a chapter can rise through a single code while the rest are unchanged, we repeat the analysis at code level.

#### 3.3.1 The Burden Carried by Rising and Falling Causes

Figure 3 gives, for each country and age group, the percentage of deaths occurring in ICD- 10 codes of each class. Codes carry fewer deaths than chapters, so a country may have several rising chapters and still few rising codes.

**Figure 3.**
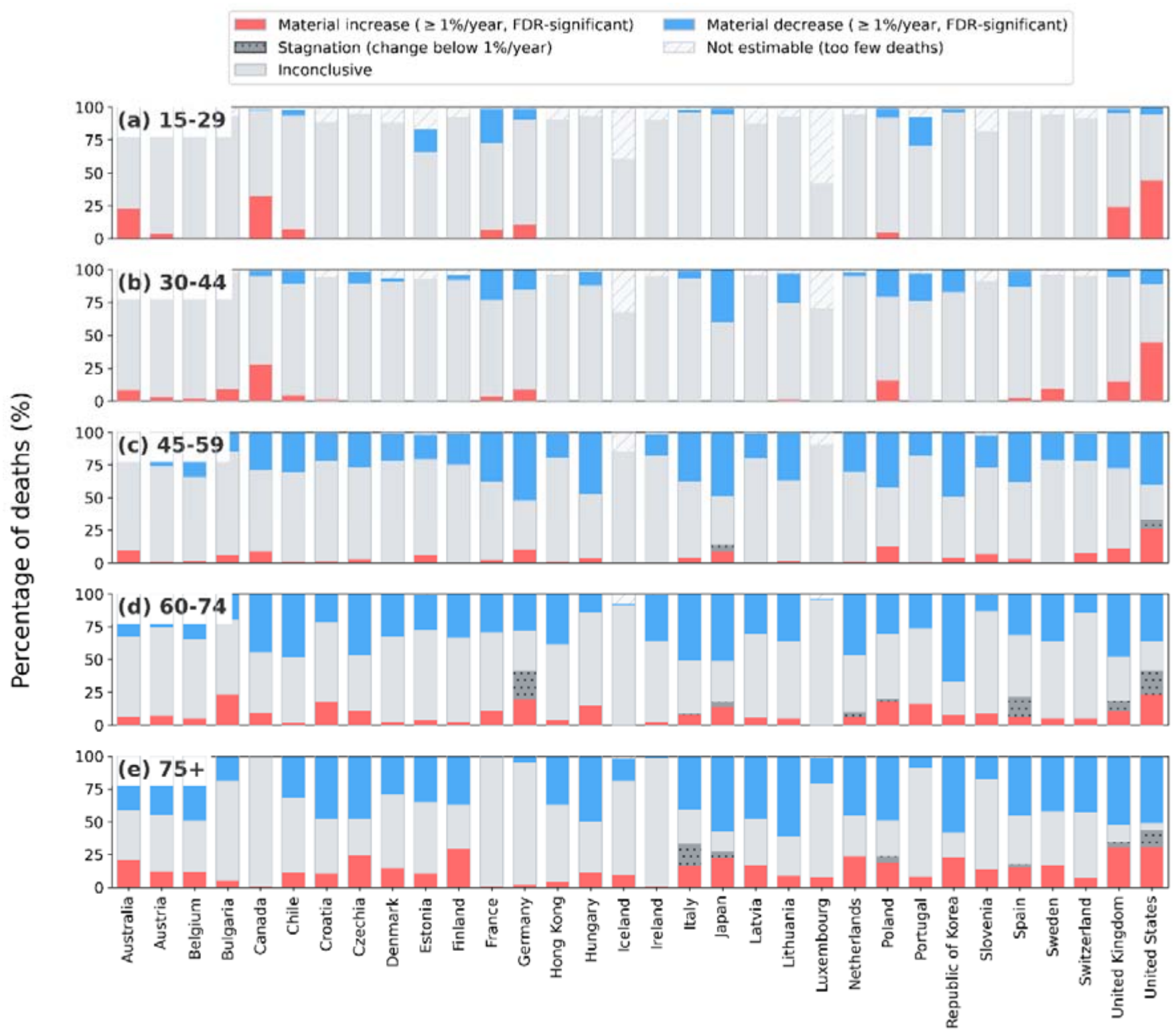
Percentage of deaths attributable to ICD-10 codes in each trend category, by country and age group. Red and blue cells indicate material changes: a rise or fall of at least 1% per year that survives Benjamini-Hochberg control of the false discovery rate. Fine stippling marks stagnation, a change below 1% per year. The unmarked pale grey is inconclusive. Diagonal hatching marks causes that could not be estimated at all. Ages 0 and 1-14 are not shown since almost no cause of death fell in a resolved trend category in any country for these ages (an average of 0.0% and 0.9% of deaths for ages 0 and 1-14, respectively).

How much of the burden can be classified at all rises steeply with age. In infancy, no cause of death in any of the 32 countries could be placed in a resolved class (increasing, decreasing, or below 1% per year), and at ages 1–14 only four countries had any such cause, accounting for 0.9% of deaths on average. The share of deaths in a resolved class averages 8.6% at ages 15–29 and 12.5% at 30–44, against 32.2% at 45–59, 43.1% at 60– 74 and 50.4% at 75 and over.

Deaths in rising causes average 4.9% at 15–29 and 4.8% at 30–44, but there are remarkable differences between countries at those younger ages. At the higher end, the US reaches 44.3% and 44.6%, Canada 32.3% and 27.7%, and the UK 24.3% and 14.9%. These shares depend on how much of a country’s mortality can be resolved, but direction does not: France and the UK resolve comparable shares at 15–29 (32.5% and 27.5%), yet 26.0 of France’s 32.5 points are declines while 24.3 of the UK’s 27.5 are increases (see Figure 3).

From 45–59 onward, the balance tips decisively toward declining causes, which rise from 27.5% of deaths on average at 45–59 to 35.3% at 75 and over while rising causes move from 4.3% to 13.7%. The countries with most of their mortality in declining causes are Germany, the Republic of Korea and Japan at 45–59 (51.7%, 49.1% and 48.6%) and Lithuania, the Republic of Korea and Japan at 75 and over (61.0%, 58.2% and 57.8%).

Exposure to rising causes at the oldest ages is widest in the US and the UK, at 30.6% and 30.4%, with Finland and Czechia close behind.

Causes whose change is below 1% per year appear only from ages 45-59 onward, and in few countries: they account for 21.7% of German and 18.4% of US deaths at 60–74, and 16.2% of Italian deaths at 75 and over, but for under 1% in most countries at every age.

#### 3.3.2 Countries most exposed to rising causes

The US carries the largest share of deaths in rising causes at every age from 15–29 onward except 60–74, where Bulgaria is fractionally ahead (23.1% against 23.0%), followed by Canada and the UK. In all three, the rising causes at 15–29 and 30–44 are injury and poisoning: intentional self-harm (X70) accounts for 14.9% of Canadian and 13.6% of UK deaths at 15–29, and accidental poisoning (X42) rises in all three at every age from 15–29 to 60–74. Assault by firearm (X95) rises in both North American countries, accounting for 10.5% of US and 1.8% of Canadian deaths at 15–29. From 45-59 onward, the three diverge. By 45–59 the leading rising codes are alcoholic liver disease (K70) and hypertensive heart disease (I11) in the US, chronic lower respiratory disease (J44) and pneumonia (J18) in the UK, and ill-defined causes (R99) in Canada. At 75 and over, dementia dominates in both the US and the UK, Alzheimer’s disease accounting for 6.6% and 4.0% of deaths and unspecified and vascular dementia adding a further 11.7% in the UK, while Canada has no rising code reaching 1% of deaths.

Two other groupings recur. In Poland, Bulgaria and Germany the rising codes are dominated by ill-defined causes and alcohol (R99, R98 and R96 alongside alcoholic liver disease and cirrhosis, K70 and K74), with heart failure alone accounting for 15.5% of Bulgarian deaths at 60–74. The dementia pattern of the oldest ages is not confined to the English-speaking countries either: in Finland, Alzheimer’s disease alone accounts for 18.4% of deaths at 75 and over and is rising.

#### 3.3.3 Causes Rising Across Many Countries

Because a small cause can rise in many countries without contributing materially to mortality, we rank codes by the number of countries in which they increase, restricted to codes accounting for at least 0.5% of deaths in the age group on average. The full ranking is in Supplementary File 2.

At ages 15–29, intentional self-harm (X70) is the most widely rising code, increasing in 4 of the 32 countries and accounting for 14.0% of deaths at that age on average. Ill-defined causes (R99) rise in 3 countries, at 4.4% of deaths.

At ages 30–44, both self-harm and ill-defined causes remain common, with R99 now the most widely rising code, in 6 countries. Accidental poisoning emerges as a prominent cause, codes X44, X41 and X42 rising in 5, 4 and 3 countries respectively.

By ages 45–59, self-harm–related causes appear less frequently among codes with increasing mortality (X70 in 2 countries), while R99 continues to feature across 9 countries. In addition, type 2 diabetes (E11), hypertensive heart disease (I11) and chronic lower respiratory disease (J44) emerge among the most frequent codes with increasing mortality, in 6, 5 and 4 countries, reflecting a shift toward chronic disease.

At 60–74, type 2 diabetes rises in 10 countries and hypertensive heart disease in 9. Neurodegenerative disease and dementia appear alongside them, with Alzheimer’s disease (G30), unspecified dementia (F03) and Parkinson’s disease (G20) each rising in 9 countries. By 75 and over, the following dominate: Parkinson’s disease and unspecified hypertensive disease (I13) each rise in 19 of the 32 countries, unspecified dementia in 18 and Alzheimer’s disease in 14, with F03 and G30 accounting for 3.9% and 3.1% of deaths at that age. R99 rises in 14 countries at 60–74 and in 17 at 75 and over, more than at any younger age.

The threshold excludes causes that rise in many countries while accounting for almost none of the deaths: influenza with virus identified (J10) rises in 25 of 32 countries at 75 and over and other degenerative diseases of the basal ganglia (G23) in 17 of 31 at 60–74, each at around 0.1% of deaths. These are more plausibly certification practice than mortality.

At code level, a sex difference was resolved in 78 of 40,000 cells, again none below age 45. Lung cancer (C34) and chronic lower respiratory disease (J44) account for 39 of the 78, with women’s rates falling more slowly in 37 of those 39.

## 4. Discussion

### 4.1 Main findings

Material increases in all-cause mortality were confined to the US and Canada at ages 15– 44, and to France in infancy, yet cause-specific increases were observed across most countries, with several continental European countries (including Poland, Germany, Italy, and Hungary) showing increases across multiple ICD-10 chapters. Stagnation in countries with already high mortality rates, such as the US, Bulgaria, Chile, and Poland, represents a further concerning pattern: in these countries, the change was estimated to be smaller than 1% per year, rather than merely unproven. Across countries experiencing increasing mortality, the drivers varied systematically by age: external causes, including drug poisoning, firearm-related deaths, and intentional self-harm, dominated in younger cohorts, while chronic conditions such as hypertensive heart disease and alcohol-related liver disease became more prominent in mid-life, and dementia and neurodegenerative disease at the oldest ages. Ill-defined causes of death recurred consistently across countries and age groups, suggesting that an important component of mortality change remains insufficiently characterised.

A central finding is that cause-specific increases are neither confined to countries with rising all-cause mortality nor concentrated at ages where all-cause mortality rises. Rising chapters became steadily more common with age, across the ages above 45 in which no country’s all-cause mortality rose. Aggregate improvement at older ages therefore conceals causes whose mortality is rising rather than falling, and a study reporting all-cause mortality alone would record a misleading uniform progress.

### 4.2 Methodological considerations

Several limitations should be noted. First, the analysis does not distinguish trends by socioeconomic characteristics such as deprivation or education; such stratification could reveal additional heterogeneity but would reduce statistical power for cause-specific analyses. The sex-specific analysis illustrates that cost directly: because the standard error of a sex difference is about twice that of a pooled trend, differences were resolved in 6.5% of country-by-age cells for all-cause mortality and only 0.2% at code level. Second, variation in cause-of-death coding, particularly for ill-defined causes, may affect cross-national comparability, although it is less likely to bias within-country trends if coding practices are stable. Third, the relatively short study period (2012–2019) may limit the detection of longer- term dynamics. Fourth, many cause-specific trends cannot be resolved at all, and we report them as inconclusive. The classification also depends on the threshold of 1% per year, but the material increases in the United States and Canada are unchanged for thresholds between 0.5% and 2% per year (Supplementary File 1, Section 6.3).

A further limitation is that ICD-10 codes capture the immediate certified cause rather than the full contribution of an exposure. Accidental poisoning (X42, X44) and alcoholic liver disease (K70) are among the rising causes reported above, yet both understate total drug- and alcohol-related mortality, which also arises from infections, organ damage and injury in which these exposures were contributory but not certified as underlying. The burden attributable to them is therefore larger than the codes imply. Finally, restricting the analysis to the pre- pandemic period does not capture the substantial mortality disruptions associated with COVID-19 or any subsequent shifts in underlying trends.

### 4.3 Interpretation in context

Our findings extend research focusing on the US and the UK^9–13,37^ to several European countries, including Poland, Germany, Italy, and Hungary, and show that the pathways differ by age: external causes at younger ages, particularly in North America^9,10^, and chronic disease in mid-life. The recurrent presence of ill-defined causes points to potential diagnostic uncertainty and limits how far cause-specific evidence can be relied upon: R99 rises in 17 of 32 countries at 75 and over, and analyses built on modelled cause hierarchies redistribute such deaths to other causes by construction^29,30^. Persistent differences between Eastern and Western Europe^2,29,38^ and continued improvement in Japan and the Republic of Korea^7,8^ underscore both enduring structural inequalities and the potential for sustained health gains.

Of the explanations advanced for these trends, austerity has the strongest cross-national evidence base, while influenza and obesity have been quantified and account for a minority of the stall^19,21–25,39^. Lacking an exposure variable or identification strategy, our design cannot test these explanations, and we treat austerity as the best-supported hypothesis rather than an established cause. Three of our results bear on it. The ages do not coincide: material all- cause increases occur only below age 45, whereas rising causes accumulate with age and are most numerous at 75 and over. The dissociation between aggregate and cause-specific trends is invisible to analyses based on life expectancy or all-cause mortality, as most of this literature is. And where it has been argued that cause-specific explanations are inherently limited because the deterioration spans many causes^18^, we find increases concentrated in a few codes recurring across many countries, dementia and neurodegenerative disease at oldest ages foremost among them.

### 4.4 Conclusions

Overall, the results suggest that stalled or worsening mortality trends during 2012-2019 reflect systemic challenges affecting multiple high-income societies rather than isolated national phenomena. The combination of rising external causes at younger ages, increasing chronic disease mortality in mid-life , dementia and neurodegenerative disease at the oldest ages, and the recurrent presence of ill-defined causes across countries points to a complex and heterogeneous set of drivers that no single national narrative can fully capture. These drivers accumulate with age even where all-cause mortality is falling, so surveillance based on all-cause rates alone would not detect them. Improving the quality and comparability of

cause-of-death data will be important for understanding these drivers and informing public health responses.

## Data Availability

The estimation code, the figure notebook and the complete output tables, with a README documenting every column, are available at https://github.com/fjpreche/Mortality_Trends_Cause.git. The analysis uses the Human Mortality Database and the WHO Mortality Database, both public and requiring free registration.

## 5. Author statement

### 5.1 Ethical approval

The presented research utilizes publicly available, aggregated and anonymized mortality and population data. No individual can be identified; therefore, ethical approval was not required.

## 6. Key Messages

- This study investigated the distribution and drivers of all-cause and cause-specific mortality trends within 32 countries between 2012 and 2019.
- While all-cause mortality increases were confined to ages under 45, cause-specific increases were widespread, with several continental European countries showing rises in multiple causes, including injuries, chronic diseases, and ill-defined conditions.
- The widespread rise of these diverse mortality drivers, including poorly understood “ill-defined” causes, indicates that stalled health progress is a systemic challenge requiring a unified public health response across high-income societies, including those where all-cause mortality continued to improve.

