## Supplementary file 1 for "Diverging Pre-Pandemic Mortality Trends: Age-Specific and Cause-Specific Patterns Across High-Income Countries"

### Supplementary File 1: mathematical formulation of the methods

Age- and cause-specific mortality trends in high-income countries, 2012–2019

**4 All-cause estimates in full** **10**

**5 Hypothesis testing multiplicity** **11**

**6 Effect size, power, and classification** **11**

#### 1 Notation and data

Let $c$ index countries, $t \in \mathcal{T} = \{t_1, \ldots, t_T\}$ the calendar years of the study period ($t_1 = 2012$, $t_T = 2019$, $T = 8$), $g$ a broad age group, and $k$ a cause of death (an ICD-10 chapter or an individual ICD-10 code). Within a broad age group, $x$ indexes a finer age unit: a single year of age for the all-cause analysis, and a World Health Organization (WHO) five-year reporting band for the cause-specific analysis. Write $\mathcal{X}_g$ for the set of finer units making up group $g$, so that $g = \bigcup_{x \in \mathcal{X}_g} x$.

The data are

$$D_{c,x,t} = \text{deaths in country } c, \text{ age unit } x, \text{ year } t, \tag{1}$$

$$E_{c,x,t} = \text{person-years of exposure}, \tag{2}$$

$$D^{(k)}_{c,x,t} = \text{deaths from cause } k. \tag{3}$$

All-cause deaths and exposures are taken from the Human Mortality Database [1] (HMD) at single years of age; cause-specific deaths are taken from the WHO Mortality Database [2] at its reporting bands and coded to the tenth revision of the International Classification of Diseases [3], with the same exposures aggregated to those bands. Cells absent from the WHO extract are genuine zeros and are set to $D^{(k)}_{c,x,t} = 0$ rather than treated as missing: discarding them would bias trends upward wherever a cause is rare.

A country was eligible if its HMD series covered all eight years $\mathcal{T}$, and if the WHO Mortality Database held ICD-10 cause-of-death data for the same eight years. HMD is the starting point for the country selection since it supplies the exposures $E_{c,x,t}$ needed for the analysis. The extract used here holds 48 series, of which seven are sub-populations of a country: East and West Germany; England and Wales, Scotland and Northern Ireland; and the New Zealand Māori and non-Māori populations. The national series is used in every case, so that the United Kingdom and Germany each enter the analysis once, and 41 countries remain.

Joint coverage of the full time window by both HMD and WHO is the binding constraint, and it excludes the nine countries listed in Table S1, leaving the 32 analysed here: Australia, Austria, Belgium, Bulgaria, Canada, Chile, Croatia, Czechia, Denmark, Estonia, Finland, France, Germany, Hong Kong, Hungary, Iceland, Ireland, Italy, Japan, Latvia, Lithuania, Luxembourg, the Netherlands, Poland, Portugal, the Republic of Korea, Slovenia, Spain, Sweden, Switzerland, the United Kingdom and the United States.

Table S1: The nine of the 41 Human Mortality Database countries that do not meet both coverage conditions. "HMD to" is the last year of the series in the extract used here; "WHO years" counts the years of 2012–2019 for which ICD-10 cause-of-death data are held. A HMD series ending before 2019 cannot supply exposures for the whole window, and fewer than eight WHO years cannot supply causes for it.

| Country | HMD to | WHO years | Condition not met |
|---|---|---|---|
| Israel | 2016 | 8 | exposures |
| Russia | 2014 | 8 | exposures |
| Belarus | 2018 | 3 | both |
| Ukraine | 2013 | 6 | both |
| Taiwan | 2023 | 0 | causes |
| Norway | 2023 | 5 (2012–2016) | causes |
| Greece | 2019 | 6 (2014–2019) | causes |
| New Zealand | 2021 | 7 (2012–2018) | causes |
| Slovakia | 2019 | 7 (2015 absent) | causes |

The age unit therefore depends on which analysis is being carried out, and with it the set $\mathcal{X}_g$ and the size of the design. Table S2 gives both.

Because the model below carries one age effect per unit, the number of parameters and the residual degrees of freedom follow directly from this table: with $n = |\mathcal{X}_g|\,T$ observations and $p = |\mathcal{X}_g| + 1$ parameters, $n - p$ ranges from 6 in the infancy band to about 250 in the top all-cause band, and is 20 for a typical mid-life chapter-level cell.

The broad groups analysed are $g \in \{0,\ 1\text{–}14,\ 15\text{–}29,\ 30\text{–}44,\ 45\text{–}59,\ 60\text{–}74,\ 75+\}$; infancy is separated from childhood because its causes of death are largely distinct, and because infants

Table S2: The age units $x$ making up each broad age group $g$. The all-cause analysis uses Human Mortality Database deaths and exposures at single years of age; the cause-specific analysis uses the WHO reporting bands, which are single years of age to 4 and five-year bands thereafter. Counts are the maximum $|\mathcal{X}_g|$: a unit with no exposure in any year is dropped, so a particular cell may have fewer. The top band is open-ended and its extent varies between countries.

| | All-cause (HMD) | | Cause-specific (WHO) | |
|---|---|---|---|---|
| Broad group $g$ | units | $\lvert\mathcal{X}_g\rvert$ | units | $\lvert\mathcal{X}_g\rvert$ |
| 0 | 0 | 1 | 0 | 1 |
| 1–14 | $1, \dots, 14$ | 14 | 1, 2, 3, 4, 5–9, 10–14 | 6 |
| 15–29 | $15, \dots, 29$ | 15 | 15–19, 20–24, 25–29 | 3 |
| 30–44 | $30, \dots, 44$ | 15 | 30–34, 35–39, 40–44 | 3 |
| 45–59 | $45, \dots, 59$ | 15 | 45–49, 50–54, 55–59 | 3 |
| 60–74 | $60, \dots, 74$ | 15 | 60–64, 65–69, 70–74 | 3 |
| 75+ | $75, \dots, \omega$ | 32–36 | 75–79, ..., 90–94, 95+ | 5 |

contribute 69% of all deaths below age 15 across the countries studied (country range 51% to 75%), so that a pooled 0–14 band estimates little more than the infant trend. For $g = 0$ the set $\mathcal{X}_g$ is a single age unit, so the age term in the model below is absent and no within-band composition effect can arise; the denominator is person-years of exposure rather than live births, so the *level* is not the conventional infant mortality rate per 1000 live births, although the annual percentage change reported here is unaffected by that choice provided it is held fixed across years. The pooled band 0–14 of the submitted version is retained for comparison only and analysed in a separate run, since as the union of 0 and 1–14 it would otherwise place overlapping tests in one family (Section 5).

### 2 Trend model

#### 2.1 Specification

A broad age group is internally heterogeneous, and a rate computed on the group as a whole is an average of its age-specific rates weighted by the observed age structure. Those weights move over the study period: within 45–59 and 60–74 the distribution shifted towards the older ages in most countries over 2012–2019, and the open-ended group 75+ is affected most of all. An aggregate rate therefore changes even when every age-specific rate within the group is constant, and a trend fitted to it confounds the two. This is the *composition effect*, and any estimate of a trend over a broad band has to deal with it.

Two routes are available. The weights can be fixed externally, by averaging the age-specific rates with those of a reference population, which is direct standardisation; or the finer age units can be modelled and the trend estimated as a parameter common to them, in which case the weights cancel of their own accord. We take the second route. It leaves the trend independent of any choice external to the data, including the choice of reference population and the correspondence between the groups analysed and the groups of a standard, and it delivers the uncertainty of the trend from the same likelihood as the trend itself — which is what later supports the minimum detectable effect of Section 6, the classification that separates demonstrated stability from absent evidence, and the sex comparison of Section 7. Section 3 reports two comparisons, different in kind: one with the unadjusted trend, which measures how much the composition effect matters and therefore whether any adjustment is needed, and one with direct standardisation, which asks whether the route taken to adjust is consistent with using a standard population.

Deaths in each (age unit, year) cell are modelled as over-dispersed Poisson counts with a log link and exposure as offset, the standard regression model for rates [6, 5]. One model is fitted per country $c$, broad age group $g$ and cause $k$:

$$D_{c,x,t}^{(k)} \sim \text{Poisson}\Big(\mu_{c,x,t}^{(k)}\Big), \qquad \log \mu_{c,x,t}^{(k)} \;=\; \alpha_{c,x}^{(k)} \;+\; \beta_{c,g}^{(k)}\,(t-\bar{t}) \;+\; \log E_{c,x,t}, \tag{4}$$

for $x \in \mathcal{X}_g$ and $t \in \mathcal{T}$, with $\bar{t} = T^{-1}\sum_{t\in\mathcal{T}} t$. The indices carry the structure of the assumption. The age effects $\alpha_{c,x}^{(k)}$ are free parameters, one per age unit (cell-means coding, no separate intercept), so the model places no constraint on the shape of the age pattern within the band. The slope $\beta_{c,g}^{(k)}$, by contrast, carries no $x$: the assumption is that within the broad group $g$ every age unit shares a common annual rate of change. The number of parameters is $p = |\mathcal{X}_g| + 1$ and the number of observations is $n = |\mathcal{X}_g| \cdot T$.

Write

$$r_{c,x,t}^{(k)} \;=\; \frac{\mu_{c,x,t}^{(k)}}{E_{c,x,t}} \;=\; \exp\Big\{\alpha_{c,x}^{(k)} + \beta_{c,g}^{(k)}(t-\bar{t})\Big\} \tag{5}$$

for the expected mortality rate in age unit $x$. Its ratio between consecutive years is $\exp\{\beta_{c,g}^{(k)}\}$ for every $x \in \mathcal{X}_g$, so the reported effect size, the *annual percentage change* of the band, is

$$\text{APC}_{c,g}^{(k)} \;=\; 100\left(e^{\beta_{c,g}^{(k)}} - 1\right) \quad \%\text{ per year}. \tag{6}$$

Because the factor $\exp\{\beta_{c,g}^{(k)}\}$ is common to every age unit in the band, any fixed weighted average of the rates (5) changes by that same factor from year to year. The estimate is therefore internally age-standardised: the composition effect is removed without a standard population being chosen [4, 5]. The price is the proportionality assumption implicit in a slope that does not depend on $x$; it is testable by adding interactions $\beta_x(t-\bar{t})$, and the shared slope is used because over an eight-year window the age-specific slopes are individually very poorly determined.

To keep the notation readable, the indices $c$, $g$ and $k$ are dropped from here on wherever a single cell is under discussion, and we write $D_{x,t}$, $\mu_{x,t}$, $\alpha_x$ and $\beta$.

#### 2.2 Estimation, over-dispersion and the covariance of $\hat{\beta}$

Collect the parameters as $\theta = (\alpha^\top, \beta)^\top$ and let $X \in \mathbb{R}^{n\times p}$ hold the age indicators alongside the centred year. The parameters are estimated by maximum likelihood, solving $X^\top(y-\mu) = 0$ with $\mu = \exp(X\theta + \log E)$ by iteratively reweighted least squares in the usual way [7].

Annual counts of deaths from a given cause are routinely more variable than Poisson. Following the quasi-likelihood approach of Wedderburn [10], the mean model is retained and the variance is scaled, $\text{Var}(D_{x,t}) = \phi\,\mu_{x,t}$, with $\phi$ estimated by the Pearson statistic

$$\hat{\phi} \;=\; \frac{1}{n-p}\sum_{x,t}\frac{(D_{x,t}-\hat{\mu}_{x,t})^2}{\hat{\mu}_{x,t}}. \tag{7}$$

Three covariance estimators are available; (Q) is the default.

$$\text{(P) Poisson:}\quad \widehat{\text{Var}}(\hat{\theta}) = \left(X^\top \hat{W} X\right)^{-1}, \tag{8}$$

$$\text{(Q) quasi-Poisson:}\quad \widehat{\text{Var}}(\hat{\theta}) = \hat{\phi}\left(X^\top \hat{W} X\right)^{-1}, \tag{9}$$

$$\text{(S) sandwich (HC1) [11]:}\quad \widehat{\text{Var}}(\hat{\theta}) = \frac{n}{n-p}\left(X^\top \hat{W} X\right)^{-1} X^\top \hat{\Omega} X \left(X^\top \hat{W} X\right)^{-1}, \tag{10}$$

with $\hat{\Omega} = \mathrm{diag}\{(D_{x,t} - \hat{\mu}_{x,t})^2\}$. Write $\mathrm{se}(\hat{\beta})$ for the square root of the corresponding diagonal element. This is the whole of the accommodation for over-dispersion. Because quasi-likelihood leaves the estimating equations for $\theta$ unchanged, $\hat{\phi}$ enters only the covariance: standard errors, intervals and the minimum detectable effect of (23) all carry it, while $\hat{\beta}$ and $\widehat{\mathrm{APC}}$ remain those of the Poisson fit.

**A floor at $\hat{\phi} = 1$.** The dispersion actually used is

$$\hat{\phi}_{\text{used}} \;=\; \max\left(\hat{\phi},\; 1\right), \tag{11}$$

so that the model never claims greater precision than Poisson. The reason is that $\hat{\phi}$ is itself estimated from $n - p$ residual degrees of freedom, which Table S2 shows can be as few as 6, and under exact Poisson sampling $\hat{\phi}$ scatters widely about 1, so a value below 1 is ordinarily sampling variation in $\hat{\phi}$ rather than evidence of genuine under-dispersion. Taking such a value at face value would shrink the standard errors on no real information. The floor binds in 17% of estimable all-cause cells, 35% at chapter level and 49% at code level, the share rising as the cells get smaller and $\hat{\phi}$ noisier, which is the pattern expected if those values are noise.

Over-dispersion is the common case in these data: $\hat{\phi}$ exceeds 1 in 83% of estimable all-cause cells (median 1.24, ninetieth percentile 2.96) and in 65% at chapter level (median 1.15, ninetieth percentile 3.47). At $\hat{\phi} = 3$ the standard errors are roughly 1.7 times their Poisson values, so the choice of variance model materially changes which trends are declared.

#### 2.3 Interval estimates and tests

Inference uses the $t$ distribution on $n - p$ degrees of freedom, which matters because the series are short. The 95% confidence interval for the APC is

$$95\%\ \text{CI for APC}: \quad 100\left(\exp\left\{\hat{\beta} \pm t_{0.975,\,n-p}\,\mathrm{se}(\hat{\beta})\right\} - 1\right), \tag{12}$$

The interval (13) is the image of the interval for $\beta$ under the strictly increasing map (6) and so has exactly its coverage. It is not symmetric about $\widehat{\mathrm{APC}}$, and its lower limit cannot fall below $-100\%$ per year. Two-sided $p$-values are obtained from the same $t$ reference distribution.

#### 2.4 Cells that are not estimable

A trend is not estimated, and no effect size is reported, when

$$\#\left\{t \in \mathcal{T} : \textstyle\sum_x D_{x,t} > 0\right\} < 3 \qquad \text{or} \qquad n \le p. \tag{13}$$

A converged fit is additionally flagged as unstable, and excluded from all classifications, when

$$|\hat{\beta}| > 3 \quad \text{or} \quad \mathrm{se}(\hat{\beta}) > 3, \tag{14}$$

corresponding to $|\mathrm{APC}|$ beyond roughly $1.9 \times 10^3\%$ per year: the signature of near-separation in a sparse series, where deaths occur only in the first or last years of the period. Such cells carry `status` $\in$ {`not estimable`, `unstable`} and are displayed as hatched, never as an absence of trend.

#### 2.5 Is a straight line adequate over the window?

Mortality improvement in high-income countries slowed at some point during the 2000s, and the timing of that slowdown has been estimated directly. McCartney et al. [12] fit segmented models to national series running from 2000 to 2019 and report a breakpoint for each country.

Restricted to the countries analysed here, 29 have an estimable breakpoint in age-standardised mortality, of which 22 (76%) fall at or before 2012, with a median of 2009.6; for life expectancy the corresponding figures are 30 countries, 21 (70%) and a median of 2008.2. Bulgaria appears in neither of their tables and Luxembourg only in the age-standardised one, and no breakpoint is identified for Chile or Hungary in age-standardised mortality. In the two countries for which we report a material mid-life increase the dated change also precedes or fall at the beginning of the window: the United States at 2010.1 in age-standardised mortality and 2011.7 in life expectancy, and Canada at 2011.8 and 2012.2.

The window 2012–2019 therefore sits after those changes for a majority of countries, which supports the use of a single slope. For the minority whose dated break falls inside the window, the slope of (4) is a period average across a change of regime.

Since the breakpoint occurs after 2012 for some countries, we run a statistical test for the linear model (4) as a null hypothesis. With $T = 8$ annual observations a segmented model must place a location parameter and two slopes, whether by the joinpoint regression common in this literature [20] or by the iterative linearisation of Muggeo [21]. Each segment would then rest on about four observations, implying that eight points cannot locate a break reliably. Accordingly, we do not estimate a break. We ask the weaker question instead: does one further degree of freedom in time improve the fit?

**The test.** Against the null model (4) the alternative

$$\mathrm{H}_1: \quad \log \mu_{x,t} \;=\; \alpha_x + \beta(t-\bar{t}) + \delta \, \max\{(t-\tau), 0\} + \log E_{x,t}, \tag{15}$$

lets the slope change once, by $\delta$, at a knot $\tau$ that is *fixed in advance* at the midpoint of the window and is not estimated. The alternative adds the single parameter $\delta$, so under quasi-likelihood the comparison is the $F$ statistic

$$F \;=\; \frac{\mathcal{D}_0 - \mathcal{D}_1}{\hat{\phi}_1} \;\sim\; F_{1,\, n-p_1} \qquad \text{under } \mathrm{H}_0, \tag{16}$$

with one numerator degree of freedom, where $p_1 = |\mathcal{X}_g| + 2$ is the number of parameters of the larger model — one level per age unit, the slope and $\delta$ — so that the denominator degrees of freedom $n - p_1$ follow the design sizes of Table S2; $\hat{\phi}_1$ is its dispersion from (7), and $\mathcal{D}$ the residual deviance $\mathcal{D} = 2\sum_{x,t}\{D_{x,t}\log(D_{x,t}/\hat{\mu}_{x,t}) - (D_{x,t} - \hat{\mu}_{x,t})\}$, with the convention $0 \log 0 = 0$.

Figure S1 gives the result for the all-cause series. A single straight line is rejected, at a false discovery rate of 5% controlled within the families of Section 5, in 34 of the 221 estimable cells, across 17 countries. For those cells the slope of (4) is a period average over a changing rate. Twenty-three of the 34 are nonetheless classified as a material change, and in 13 the slopes on either side of the knot have opposite signs. Rejection is evidence that a straight line is inadequate for a series; failure to reject is not evidence that it is adequate, since the test has limited power where deaths are few.

**Comparison with the dated breakpoints.** Set against McCartney et al.'s dating [12], the two analyses agree in two distinct ways.

Of the 12 countries whose dated breakpoint falls after 2012 on either of their measures, our test rejects a straight line in 8: Canada, Croatia, Czechia, Denmark, Finland, Poland, Spain and the United Kingdom. Where a change of regime has been dated inside our window, we tend to detect it. Of the 19 whose dated breakpoints both precede 2012, our test still rejects in 8: Australia, Ireland, Japan, Latvia, Lithuania, Portugal, the Republic of Korea and the United States. In these the rate of improvement changed again within 2012–2019, after the change they date. Bulgaria, absent from both of their tables, rejects at ages 45–59.

The two analyses also differ in resolution, which is why a country can appear in either group. The breakpoints tabulated by McCartney et al. are fitted to all-age series, one date per country, whereas our test is applied within each of seven age groups, so a country may reject at some ages and not others: Poland rejects in five of its seven age groups, while Canada and the United Kingdom reject in one each, at ages 60–74 and 75+ respectively, away from the mid-life estimates on which our conclusions rest. McCartney et al. do fit segmented models by age group, but with coarser bands above age 30 and without tabulating the resulting dates, so an age-by-age comparison is not available.

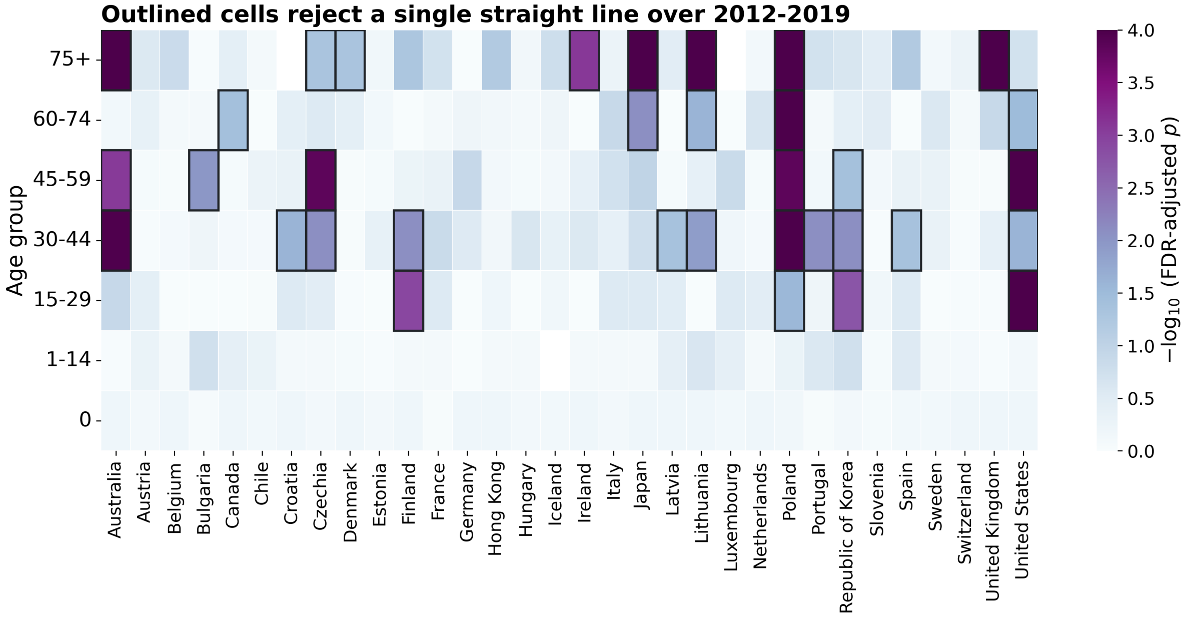

Figure S1: Adequacy of a single log-linear trend over 2012–2019, all-cause series. Colour is the false discovery rate adjusted $p$-value of the fixed-knot test of (17), on a $-\log_{10}$ scale, by country and age group. Outlined cells are those rejecting a straight line; for them the slope of (4) is a period average across a change of regime. Failure to reject does not establish linearity, since the test has limited power where deaths are few.

### 3 Conventional estimators and mortality levels

Two further trends are computed for every all-cause series and published with the estimate tables, as diagnostics rather than as results. The first makes no adjustment for age structure at all, and so measures how much the adjustment matters; the second adjusts by the other of the two routes of Section 2, and so asks whether the route taken changes the answer. Figure S2 shows a larger difference in the first comparison than in the second, supporting the approach taken here. The standardised rate of Section 3.2 then serves a second purpose, reported in Section 3.3: it gives the mortality level of each country, which the trend alone does not.

#### 3.1 The unadjusted trend

The first comparison is with the trend of the band's overall rate: all deaths in the band divided by all person-years in the band, drawing no distinction between the ages inside it. Operationally this is (4) fitted to the aggregated series $(\sum_x D^{(k)}_{c,x,t},\ \sum_x E_{c,x,t})$, with the age term $\alpha^{(k)}_{c,x}$ absent because the band is then a single age unit. Taking the age structure as it comes is what exposes this estimator to the drift in that structure described in Section 2, so its difference from the primary estimate is a direct measure of the composition effect, in percentage points per year.

The left panel of Figure S2 shows that difference growing with age: it is zero in infancy, where the band is a single age unit and the two estimators coincide by construction, and reaches a median of about 0.7 percentage points per year at ages 75 and over, where in four countries the two disagree about the direction of travel. This highlights the importance of adjusting for the age structure within a broad band.

The sharpest single case is the United Kingdom at ages 45–59. The unadjusted trend is an increase of 0.43% per year; with the age term restored the estimate is a decline of 0.05% per year, with a confidence interval spanning zero. The sign of the trend in that band is therefore a property of the within-band age structure rather than of the mortality rates, and the same adjustment works in the other direction at ages 30–44, where it raises the United Kingdom estimate from 0.18% to 0.62% per year and makes an increase understated by crude rates distinguishable from zero. Both estimates are reported for every cell, so the size of the effect can be read directly.

##### 3.2 Direct standardisation

The second comparison is with the conventional remedy, direct standardisation to the European Standard Population 2013 [16]. The standard is defined on five-year age groups, which are coarser than the single years of age used by the model for the all-cause analysis, so the strata of this estimator are the standard's own groups. Let $\mathcal{J}_g$ be the set of them lying inside the broad group $g$, let $W_j$ be the standard population of group $j$, and let

$$w_j \;=\; \frac{W_j}{\sum_{j' \in \mathcal{J}_g} W_{j'}}, \qquad j \in \mathcal{J}_g, \qquad \sum_{j \in \mathcal{J}_g} w_j = 1, \tag{17}$$

be those populations renormalised over the broad group. With $D^{(k)}_{c,j,t} = \sum_{x \in j} D^{(k)}_{c,x,t}$ and $E_{c,j,t} = \sum_{x \in j} E_{c,x,t}$ the deaths and exposures of stratum $j$, the directly standardised rate of the broad group is the weighted average of the observed stratum rates,

$$R^{(k)}_{c,g,t} \;=\; \frac{1}{\sum_{j \in \mathcal{J}^+_{g,c}} w_j} \sum_{j \in \mathcal{J}^+_{g,c}} w_j \, \frac{D^{(k)}_{c,j,t}}{E_{c,j,t}}, \qquad \mathcal{J}^+_{g,c} = \{ j \in \mathcal{J}_g : E_{c,j,t} > 0 \}, \tag{18}$$

the leading factor restoring unit total weight when a stratum is absent from a country's exposure series. The trend is then taken from the eight annual values of (20) by ordinary least squares on their logarithm,

$$\tilde{\beta}^{(k)}_{c,g} \;=\; \frac{\sum_{t \in \mathcal{T}} (t - \bar{t}) \, \log R^{(k)}_{c,g,t}}{\sum_{t \in \mathcal{T}} (t - \bar{t})^2}, \qquad \widetilde{\mathrm{APC}}^{(k)}_{c,g} \;=\; 100 \left( e^{\tilde{\beta}^{(k)}_{c,g}} - 1 \right), \tag{19}$$

on the same scale as (6) and so directly comparable with it.

Two features of (20) deserve note, because the referee's concern about the fit between the standard and the data bears on both. The broad groups of Table S2 coincide with the boundaries of the standard's five-year groups, so each $\mathcal{J}_g$ consists of whole standard groups and the $W_j$ of (19) are never split between broad groups. And the standard's open-ended group, ages 95 and over, is matched to the open-ended top of the exposures rather than closed at an arbitrary age, so the oldest ages enter (20) with the weight the standard assigns them instead of one determined by where a country's exposure series happens to stop. Had it instead been closed at the top of the exposure range and its weight apportioned pro rata over single years of age, the weight carried by ages 95 and over within 75+ would have fallen from 2.2% to about 0.3%, and the standardised rate of the oldest band would have been dominated by ages 75–84.

The right panel of Figure S2 shows none of the structure of the left: standardising to an external population reproduces the primary estimate almost exactly, with a median absolute difference of 0.05 percentage points per year. The two routes agree on the trend, and the difference between them lies elsewhere. Standardising first and regressing the resulting eight annual values afterwards gives a slope whose standard error reflects only the scatter of those eight summaries, taking no account of how many deaths lie behind each, whereas the interval of (13) is built from the counts and carries the over-dispersion of Section 2.2. What the model adds over direct standardisation is therefore not a different trend but an interval for it.

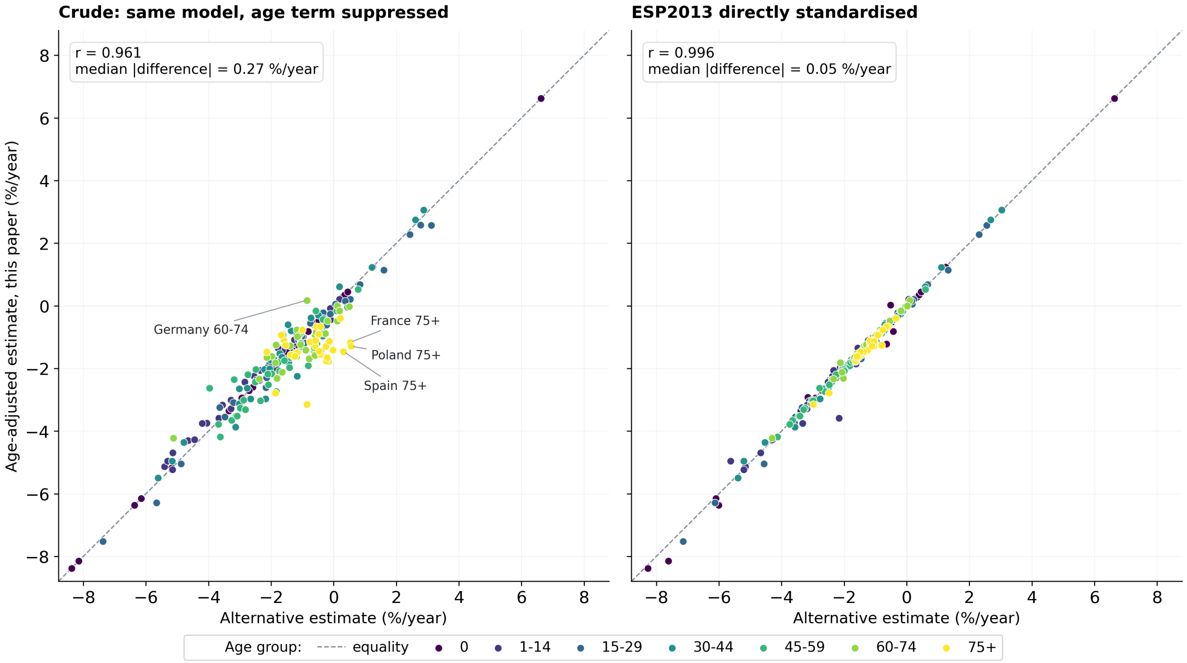

Figure S2: The primary estimate against the two quantities computed alongside it, for every estimable all-cause series. The two panels answer different questions. Left: the unadjusted trend, the same model with the age term suppressed, which shows how much the age structure matters. Right: the ordinary least squares slope of the log ESP2013 directly standardised rate, a second way of adjusting for it. The dashed line is equality: departures from it grow with age in the left panel and are absent in the right.

#### 3.3 Mortality levels

The trends reported in the main text say how fast mortality is moving, not where a country starts, and some statements in the Discussion rest on the level as well as on the trend. The standardised rate (20) supplies it, and the argument for standardising is the one just made: a crude rate over a broad band reflects how a country's population is distributed inside that band as well as its mortality, so comparing levels across countries without standardising would reintroduce the confounding that Section 2 removes from the trend.

The level reported is (20) at the middle of the window. Under the log-linear trend of (4) the fitted mid-point is the geometric mean of the first and last year, so no further fitting is required. Rates differ by three orders of magnitude between the youngest and the oldest band, so Figure S3 shows each country as a ratio to the median country of its own age group, on a logarithmic scale, with the median of each row given in the row label. Levels are reported for every cell with deaths and exposure, including those few whose trend could not be estimated stably: instability is a property of the slope, not of the rate.

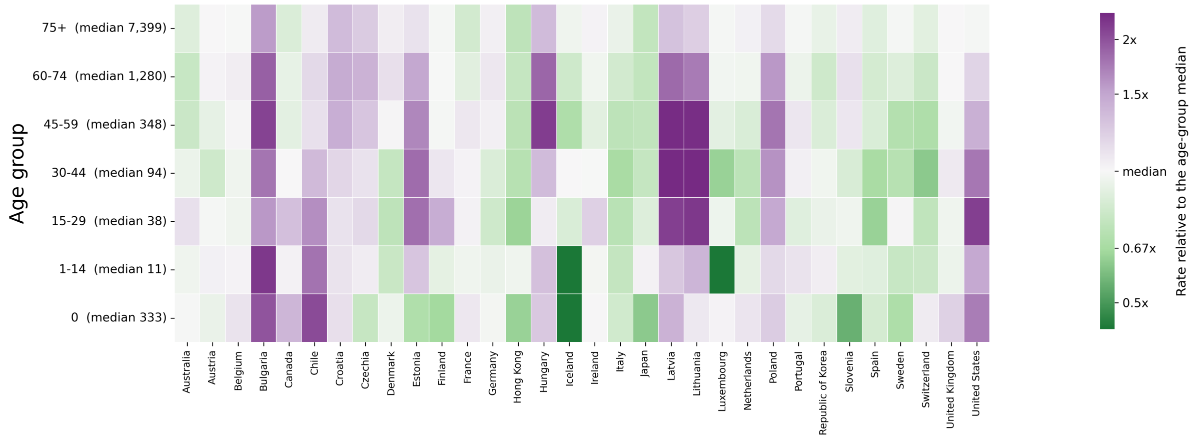

Figure S3: Mortality level by country and age group, 2012–2019. Colour is each country's directly standardised rate at mid-period as a ratio to the median country of the same age group, on a logarithmic scale; the median rate of each age group, per 100 000 person-years, is given in the row label. The colours are deliberately not those of the trend maps, where red and blue mean increasing and decreasing. Hatched cells have no rate.

### 4 All-cause estimates in full

The all-cause results are shown in Figure 1 of the main text, which gives the estimated annual percentage change for every country and age group. Figure S4 gives every age group in that form, so no age group is represented only by a colour. One country order, taken from the ages 30–44 estimates, is shared by all seven panels: a country keeps its row throughout, so a pattern running across several age groups can be read down the figure, as in the colour map, while each estimate still carries its interval.

Read with Figure S3, these panels separate two things that the colour map necessarily merges: how precisely a country's trend is known, which is the width of its interval, and how far that trend is from zero. Countries whose interval spans the shaded band of ±1% per year are classified as inconclusive rather than stable, because the deaths available cannot tell the two apart.

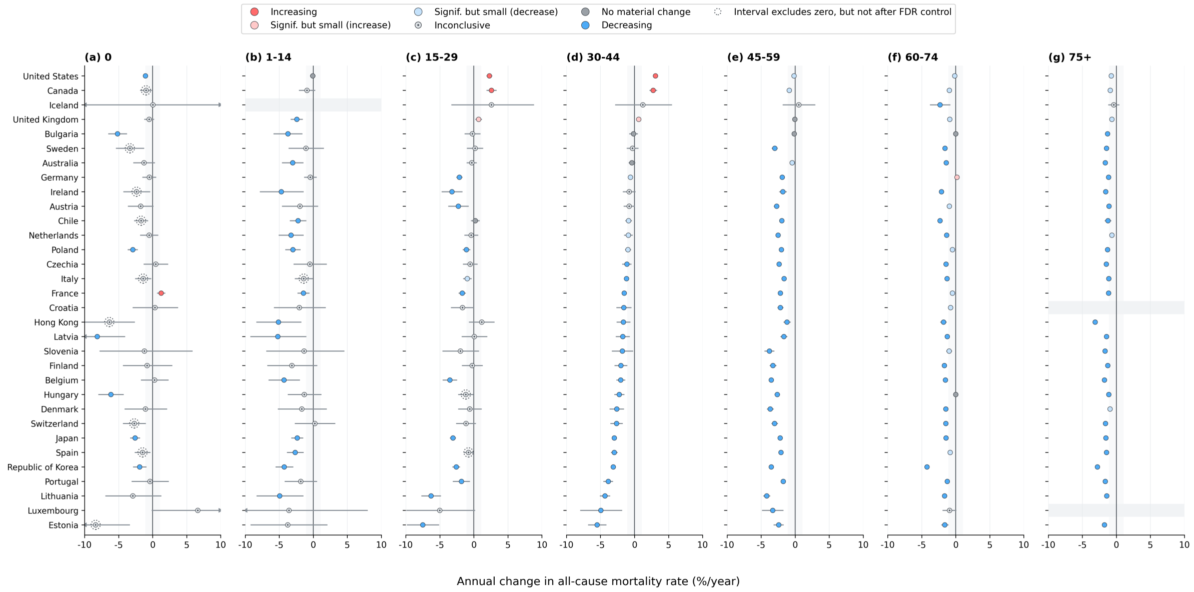

Figure S4: Annual percentage change in all-cause mortality with 95% confidence intervals, by country and age group. The country order is the same in every panel, taken from the ages 30–44 estimates, and all panels share one horizontal scale; intervals reaching beyond it are drawn with arrows. Marker colour follows the classification of Section 6 and the shaded band marks changes of less than 1% per year; intervals are unadjusted, whereas the classification uses adjusted $p$-values, so an interval may exclude zero without the country being classified as increasing or decreasing, and markers ringed with a dotted circle are those cases. A faint band marks a cell with no estimate.

### 5 Hypothesis testing multiplicity

The analysis performs one test per (country, age group, cause) cell, which amounts to $\sim 10^4$ tests at code level. Let $\mathcal{F}$ denote a declared *family* of tests, taken here to be all country $\times$ cause tests sharing a sex, age group and cause level, and let $p_{(1)} \leq \cdots \leq p_{(m)}$ be the ordered $p$-values within $\mathcal{F}$. The Benjamini–Hochberg [17] adjusted values are

$$q_{(i)} \;=\; \min_{j \geq i} \; \min\left\{1,\; \frac{m}{j}\, p_{(j)}\right\}, \tag{20}$$

which control the false discovery rate at level $q$ under independence or positive regression dependence. Trends for related causes within a country are positively dependent, so (22) is appropriate. Cells of Section 2.4 contribute no $p$-value and do not enter $m$.

Controlling the false discovery rate addresses how many of the declared discoveries are spurious, not whether a discovery is large enough to matter; the remedy is to report the magnitude of the effect with an interval rather than a dichotomy [18], which is the question addressed next.

### 6 Effect size, power, and classification

#### 6.1 Minimum detectable effect

For a two-sided test at level $\alpha$ with power $1 - \zeta$, and using the normal approximation to the distribution of $\hat{\beta}$ that underlies the standard sample-size calculations for Poisson regression [19], the smallest slope that would be declared significant satisfies $|\beta| \geq (z_{1-\alpha/2} + z_{1-\zeta})\,\mathrm{se}(\hat{\beta})$. With

$\alpha = 0.05$ and $1 - \zeta = 0.8$, the minimum detectable annual percentage change is

$$\mathrm{MDE}_{80} \;=\; 100\left(\exp\left\{2.8016 \times \mathrm{se}(\hat{\beta})\right\} - 1\right). \tag{21}$$

(The normal quantiles are used in (23) even though tests use the $t$ distribution; this understates the required effect slightly and is reported as a planning quantity, not as an inferential one.) Reporting $\mathrm{MDE}_{80}$ for every cell distinguishes a country whose mortality is demonstrably flat from one in which no trend of practical size could have been detected. Figure S5 gives it for the all-cause analysis, and Figure S6 shows the same constraint from the other direction: across every cause and age group, the estimates that are large are the ones whose standard errors are large, so that magnitude and imprecision travel together.

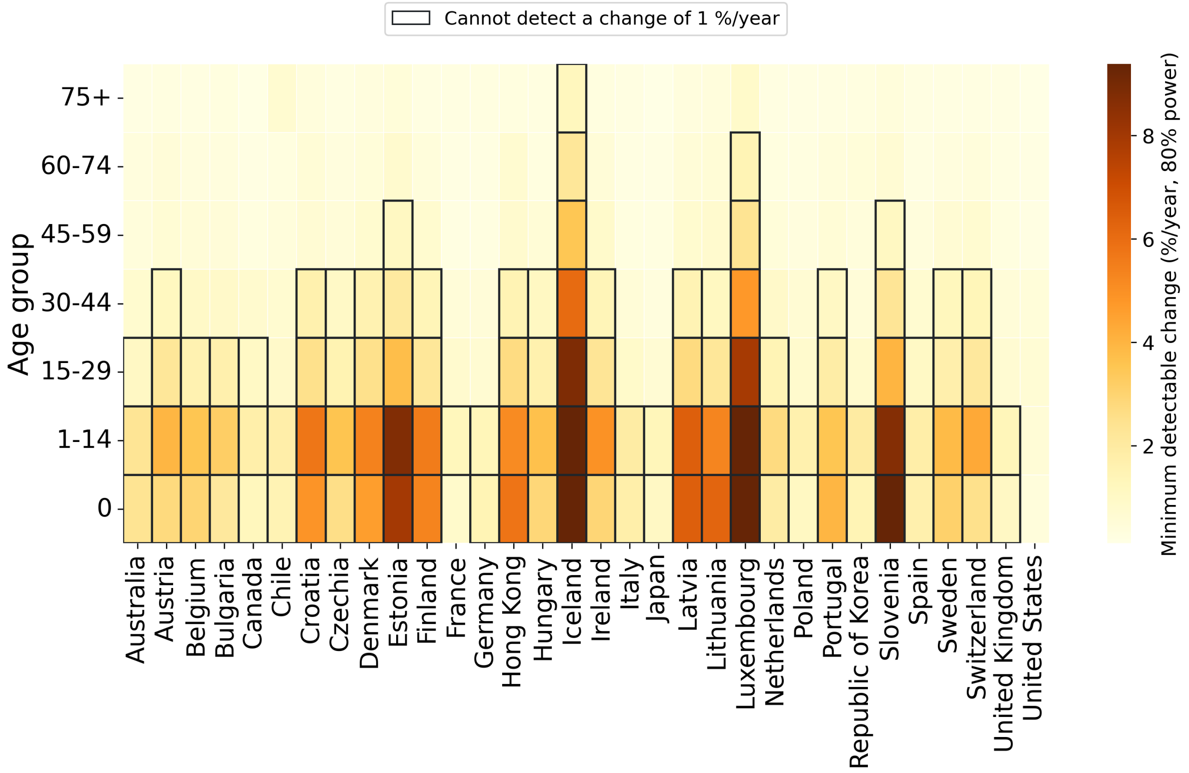

Figure S5: Smallest annual change detectable with 80% power at a two-sided 5% level, by country and age group, for the all-cause analysis. Outlined cells could not have detected a change of 1% per year, the threshold used in the classification of Section 6.2; in those cells the absence of a detected trend is not evidence of stability, whereas a trend that is detected there is one large enough to have cleared a wide interval.

#### 6.2 Classification of estimable cells

Fix a materiality threshold $\delta$ (the analysis uses $\delta = 1\%$ per year). With $q$ the adjusted $p$-value of (22) and $(L, U)$ the confidence limits of (13), each estimable cell is assigned

$$\text{category} = \begin{cases} \text{increasing}, & q < 0.05,\ \widehat{\mathrm{APC}} \geq \delta, \\ \text{decreasing}, & q < 0.05,\ \widehat{\mathrm{APC}} \leq -\delta, \\ \text{significant but small}, & q < 0.05,\ |\widehat{\mathrm{APC}}| < \delta, \\ \text{no material change}, & q \geq 0.05,\ (L, U) \subset (-\delta, \delta), \\ \text{inconclusive}, & q \geq 0.05,\ (L, U) \not\subset (-\delta, \delta). \end{cases} \tag{22}$$

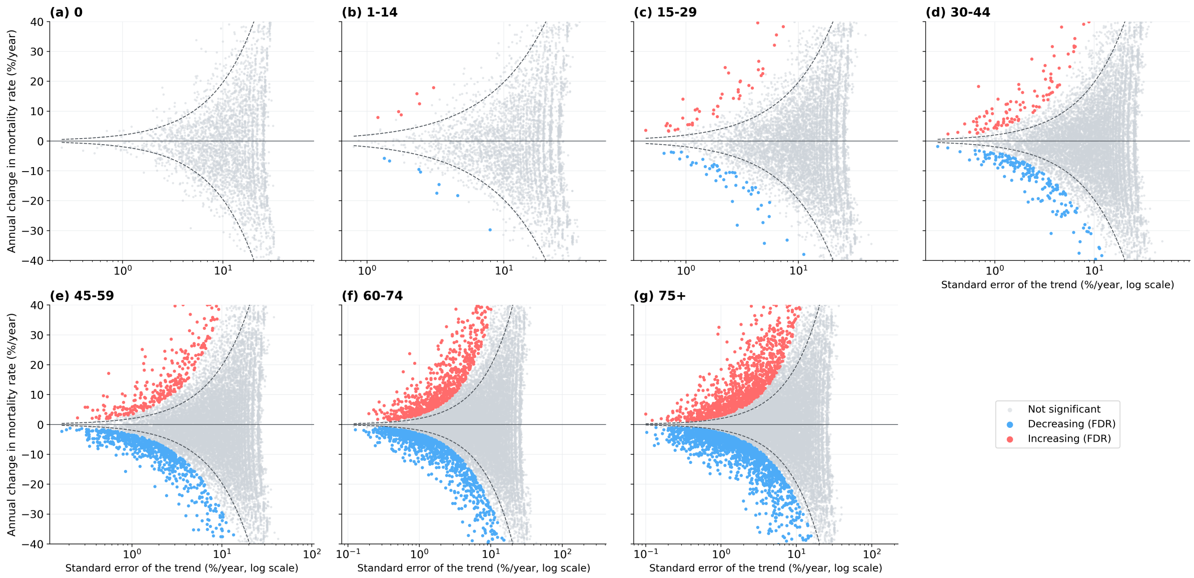

Figure S6: Estimated annual change against its standard error at ICD code level, one panel per age group, over every country and cause. Dashed lines mark the nominal two-sided 5% boundary; grey points beyond them are nominally significant but did not survive control of the false discovery rate of Section 5. Estimates that are large are predominantly those whose standard errors are large, so a large point estimate in a small cell is not evidence of a correspondingly large trend.

The first three cases are exhaustive when $q < 0.05$, so the last two apply exactly when the trend is not distinguishable from zero after adjustment. They say different things. The fourth is an equivalence statement: the whole interval lies inside $(-\delta, \delta)$, so a change of material size is excluded in either direction. The fifth covers cells whose interval still admits one, which is absence of evidence rather than evidence of absence; separating the two keeps a cell with too few deaths to resolve a trend from being read as a cell with no trend.

Figure 1 of the main text displays these classes with four marks, colour having already separated an increase from a decrease. A filled dot marks the first two classes together, a material change. An open circle marks the third, a change that survives the same control but is estimated below $\delta$. Cross-hatching marks the fourth, in which no change was detected and one of material size is excluded. A cell carrying none of these marks is in the fifth class, where the deaths available cannot resolve a change of that size. The third and fourth are reported together as stagnation, which is a claim about magnitude rather than about significance; the figure keeps them apart because the evidence behind them differs. Of the 221 estimable all-cause cells, 126 carry a filled dot, 25 an open circle, 8 are cross-hatched and 62 are unmarked. Diagonal hatching marks cells that could not be estimated (Section 2.4).

#### 6.3 Choice of the threshold

The threshold $\delta$ states what size of change is worth reporting. It is set at 1% per year and applied unchanged to every cell, so that a trend called material means the same thing at every age, in every country and at every level of cause. Its size is of the order of the improvement these populations were achieving anyway: the median all-cause trend over 2012–2019 is a decline of between 1.0% and 2.4% per year depending on age group, so a departure of 1% per year is a material fraction of the prevailing rate of progress.

What the data can resolve is a separate matter, set out in Section 6.1. At all-cause level the median minimum detectable change is close to $\delta$, so about half of the cells can tell a material change from none; at chapter and code level it is several times larger, which is why so many cause-specific cells are reported as inconclusive.

Table S3 repeats the all-cause classification at other thresholds.

Table S3: All-cause classification of the 221 estimable cells under other materiality thresholds. Stagnation pools the third and fourth classes of (24).

| $\delta$ (% per year) | Increasing | Decreasing | Stagnation | Inconclusive |
|---|---|---|---|---|
| 0.5 | 7 | 138 | 11 | 65 |
| 1.0 | 5 | 121 | 33 | 62 |
| 2.0 | 4 | 64 | 109 | 44 |

The conclusions about increases do not turn on the choice. The same four cells — the United States and Canada at ages 15–29 and 30–44 — are material increases at every threshold from 0.5% to 2% per year, and no increase from age 45 onwards is material at any of them. The remaining increases lie nearer the threshold: France's infant increase of 1.24% per year is material at $\delta = 1\%$ but not at 1.25% or above, and the United Kingdom's mid-life increases, 0.68% at ages 15–29 and 0.62% at 30–44, count as increases only at a threshold of 0.6% or less. The numbers of material decreases and of stagnating cells do depend on $\delta$, as they must: a higher threshold moves cells into stagnation from both the material and the inconclusive classes, and changes the ordering of countries within each.

##### 6.4 Aggregate measures

For each country and age group, the share of deaths falling in each category is

$$S_{c,g}(\text{cat}) \;=\; 100 \times \frac{\sum_{k\,:\,\text{cat}(k)=\text{cat}} \; \sum_t D^{(k)}_{c,g,t}}{\sum_k \; \sum_t D^{(k)}_{c,g,t}}, \tag{23}$$

and the absolute counterpart to (6), reported alongside it because a large relative change in a rare cause is of little public-health consequence, is the ordinary least squares slope of the crude cause-specific rate, in deaths per $10^5$ per year.

#### 7 Sex-specific analysis

##### 7.1 Model

Both sources carry sex, so the extension requires no new data. Let $s \in \{0, 1\}$ index sex ($s = 1$ for males, say) and extend (4) to

$$\log \mu_{x,s,t} \;=\; \alpha_{x,s} \;+\; \beta\,(t-\bar{t}) \;+\; \gamma\, s\,(t-\bar{t}) \;+\; \log E_{x,s,t}. \tag{24}$$

The intercepts $\alpha_{x,s}$ are fully crossed in age and sex, so the two sexes are allowed completely different age patterns of mortality; only the *trends* are compared. The estimands are

$$\text{APC}_{\text{female}} = 100\left(e^{\beta} - 1\right), \qquad \text{APC}_{\text{male}} = 100\left(e^{\beta+\gamma} - 1\right), \tag{25}$$

$$\text{ratio of annual rate ratios} \quad \Gamma = e^{\gamma}, \qquad \text{difference in trends} \quad \approx 100\,\gamma \;\; \text{pp/year}. \tag{26}$$

The null hypothesis of interest is $H_0 : \gamma = 0$ — the trends are parallel on the log scale — tested by the Wald statistic $\hat{\gamma}/\text{se}(\hat{\gamma})$ against $t_{n-p}$, with $n = 2|\mathcal{X}_g|T$ and $p = 2|\mathcal{X}_g| + 2$.

Three points favour (26) over fitting the two sexes separately. First, the contrast $\beta + \frac{1}{2}\gamma$ is the trend of a rate standardised to an equal mix of the sexes, and in the balanced case its variance equals that of the pooled slope of (4), so no precision is lost by allowing the trends to differ (this is a sex-standardised quantity, not the crude trend of the combined population, from which it differs whenever the sex composition of the group changes over time). Second, the comparison is made by a single test with an interval on the difference, rather than by comparing two significance verdicts — the error of declaring a sex difference because one sex reached significance and the other did not. Third, it halves the number of tests entering the family $\mathcal{F}$ of Section 5.

##### 7.2 Multiplicity for the interaction

The interaction tests form their own family: one test per (country, age group, cause) cell, with (22) applied within sex-independent strata of age group and cause level. Because this family is half the size of the sex-stratified alternative, and because $\gamma$ is the quantity of scientific interest, the recommended reporting is the pooled trend and its interval, the two sex-specific trends and their intervals, and $\hat{\gamma}$ with its interval and adjusted $p$-value — with sex differences claimed only where the last is significant.

##### 7.3 Results of the sex-specific analysis

Figures S7 to S9 give the sex-specific results.

For all-case mortality, resolved differences are confined to the middle of the age range (Figures S7 and S8). An absence of marked differences at the extremes is not evidence that the sexes moved together.

At chapter level, this is pronounced enough to govern what Figure S9 can show. Of the 2,834 estimable country-by-chapter cells, 56 (2.0%) carry a sex difference that is both significant after false discovery rate control and at least $\delta = 1$ percentage point per year, and every one of them lies at ages 45–59 or above: 15 at 45–59, 19 at 60–74 and 22 at 75+. Below age 45 not one of the 1,387 estimable cells — ages 0, 1–14, 15–29 and 30–44 — meets both conditions. The figure therefore shows only the three oldest age groups, and within them only the chapters containing at least one such cell. The contrast with the all-cause analysis is instructive: there, resolved differences appear from age 30–44 onwards, because pooling causes buys the precision that the chapter split gives away.

The same models were fitted for individual ICD-10 codes. Of the 40,000 estimable country-by-code cells, 78 (0.20%) carry a resolved difference, and again none lies below age 45: 10 at 45–59, 25 at 60–74 and 43 at 75+. The progression across the three levels of aggregation is the clearest statement in this supplement of what subdividing causes costs:

| Level | Estimable cells | Resolved differences | Share |
|---|---|---|---|
| All causes combined | 216 | 14 | 6.5% |
| ICD-10 chapter | 2,834 | 56 | 2.0% |
| ICD-10 code | 40,000 | 78 | 0.20% |

Two features of the resolved code-level differences are worth recording. First, they concentrate in a small number of causes: malignant neoplasm of the bronchus and lung (C34) accounts for 24 of the 78 and chronic lower respiratory disease (J44) for a further 15, so two codes carry half of them. Second, the direction is consistent — in 54 of the 78 it is the female rate that fell more slowly: in all 24 of the C34 cells and in 13 of the 15 J44 cells, the exceptions being Hong Kong and Japan at 75+. The resolved differences are listed in full in the `Code-level differences` sheet of Supplementary File 2.

Applying the classification of Section 6 to the difference rather than to a trend, a further 35 cells (1.2%) fall in the two stagnation classes, the difference being either significant but estimated below $\delta$ or non-significant with an interval lying entirely inside $(-\delta, \delta)$; the remaining 2,743 cells (96.8%) are unresolved. Because so few cells can be placed on either side of $\delta$, the marks used in Figure 1 of the main text for those two classes are not reproduced in Figure S9: together they would apply to 35 cells in 2,834. Deaths before midlife are few, and splitting them further by sex leaves even less to resolve a difference with. The absence of resolved sex differences below age 45 is thus a statement about what these data can establish, not a demonstration that men and women moved together.

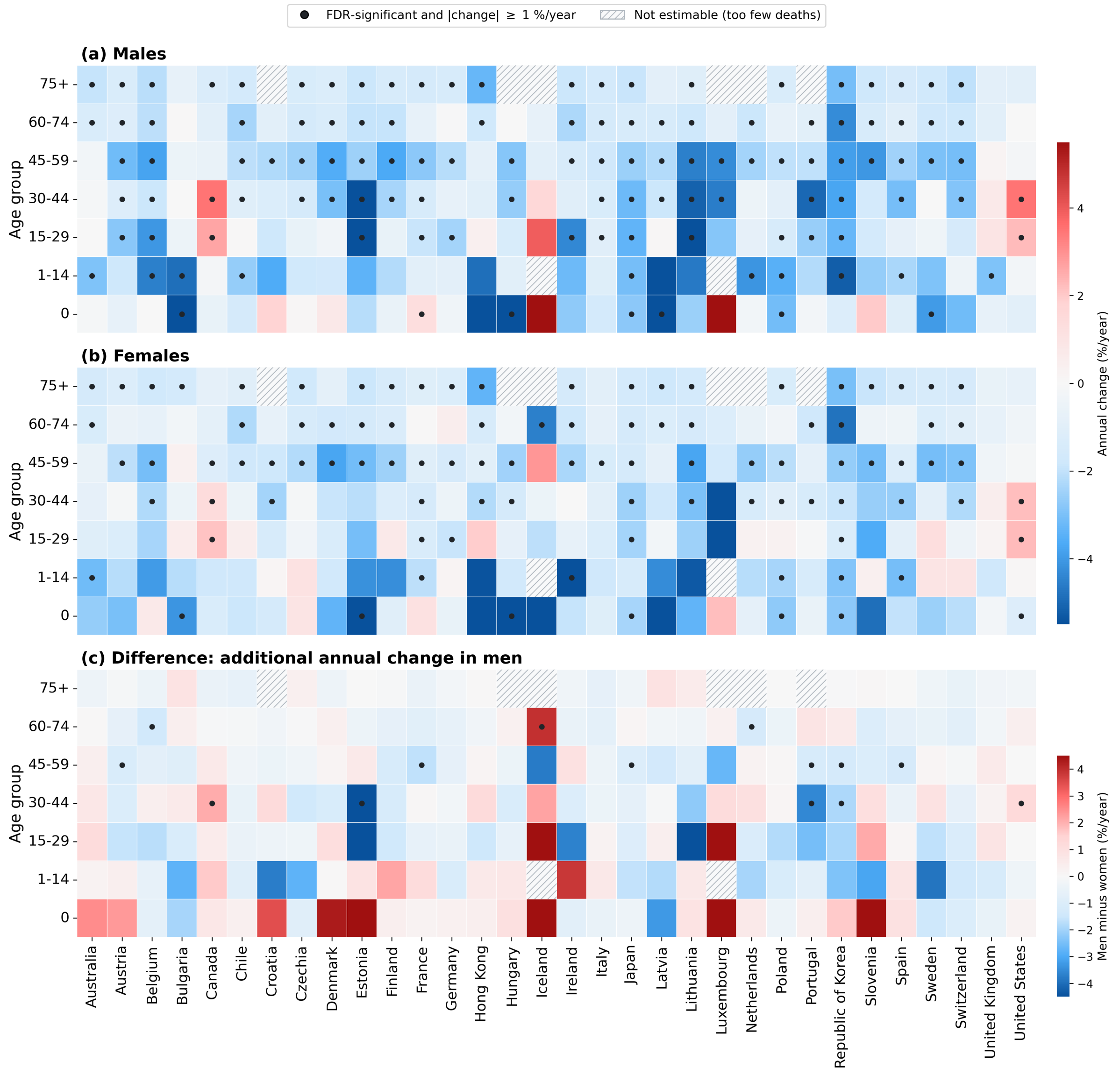

Figure S7: Age-by-sex trends in all-cause mortality, 2012–2019, by country and age group: (a) men, (b) women, (c) the additional annual percentage change in men relative to women, estimated as the sex-by-year interaction $\gamma$ of (26), one model per cell. Dots mark estimates surviving Benjamini–Hochberg control of the false discovery rate, applied within each test set separately, that also exceed 1% per year in magnitude. Hatched cells could not be estimated.

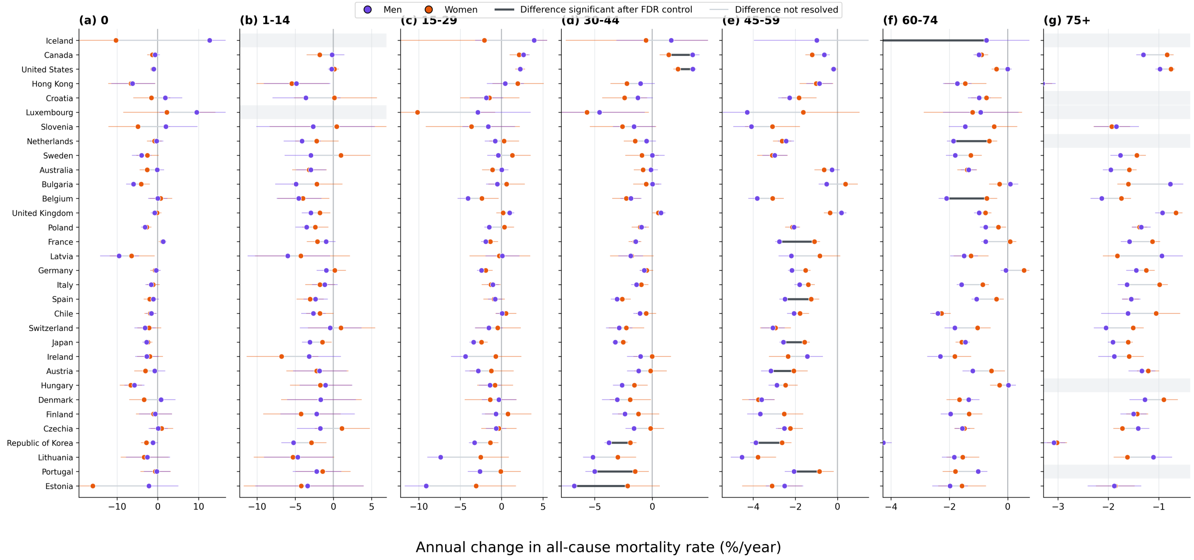

Figure S8: Annual percentage change in all-cause mortality for men and women with 95% confidence intervals, by country and age group. The bar joins the two estimates, and is darker and thicker where the difference survives false discovery rate control. One country order, taken from the difference at ages 30–44, is shared by every panel, so a country keeps its row across age groups and a pattern spanning several ages can be read down the figure; the names are given once, on the first panel, and a faint band marks a cell with no estimate. The horizontal scale does differ between panels: the plotted range runs from about −21 to +19% per year in infancy but only from −2.6 to −0.4% per year at ages 75 and over, so a common scale would compress the older panels to a few pixels. A few very wide intervals are truncated at the axis.

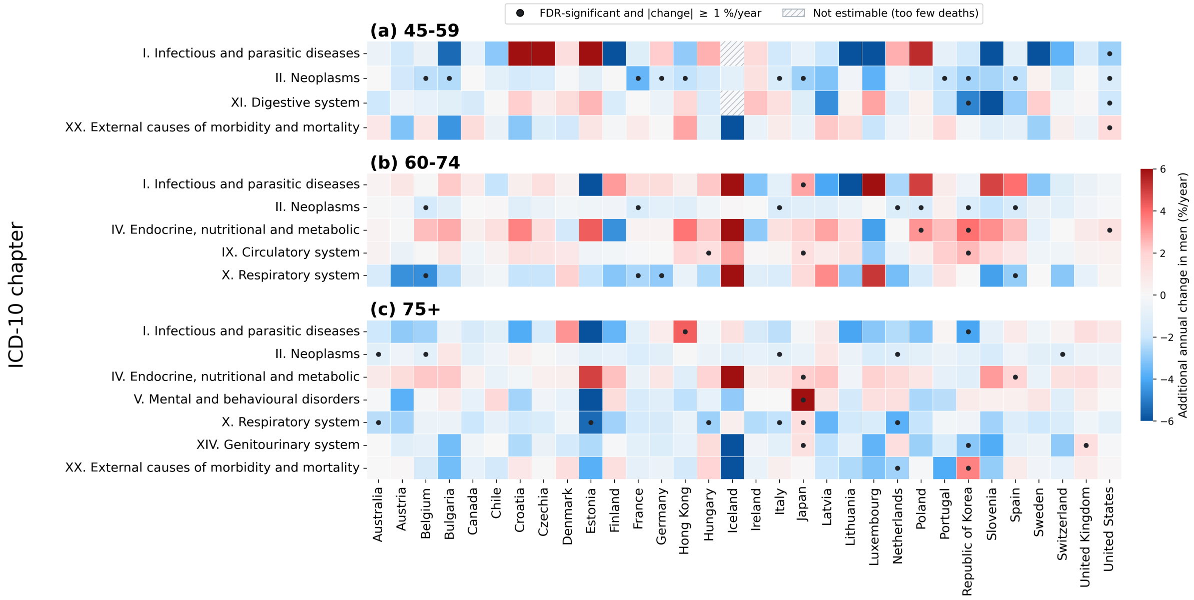

Figure S9: Additional annual percentage change in men relative to women by ICD-10 chapter and country. Only the age groups in which at least one chapter-by-country cell showed a sex difference that was both significant after false discovery rate control and at least $\delta = 1$ percentage point per year are shown, and within them only the chapters containing such a cell; in ages 0, 1–14, 15–29 and 30–44 no cell met both conditions. Details as Figure S7.

#### References

[1] Human Mortality Database. University of California, Berkeley, and Max Planck Institute for Demographic Research. Available at `https://www.mortality.org`.

[2] World Health Organization. *WHO Mortality Database.* Geneva: World Health Organization. Available at `https://www.who.int/data/data-collection-tools/who-mortality-database`.

[3] World Health Organization. *International Statistical Classification of Diseases and Related Health Problems*, 10th revision, 5th edn. Geneva: World Health Organization, 2016.

[4] Breslow NE, Day NE. *Statistical Methods in Cancer Research. Volume II: The Design and Analysis of Cohort Studies.* IARC Scientific Publications No. 82. Lyon: International Agency for Research on Cancer, 1987.

[5] Clayton D, Hills M. *Statistical Models in Epidemiology.* Oxford: Oxford University Press, 1993.

[6] Frome EL. The analysis of rates using Poisson regression models. *Biometrics* 1983;39:665–74.

[7] McCullagh P, Nelder JA. *Generalized Linear Models*, 2nd edn. London: Chapman and Hall, 1989.

[8] Penrose R. A generalized inverse for matrices. *Math Proc Cambridge Philos Soc* 1955;51:406–13.

[9] Rao CR, Mitra SK. *Generalized Inverse of Matrices and Its Applications.* New York: Wiley, 1971.

[16] Eurostat. *Revision of the European Standard Population: report of Eurostat's Task Force.* Luxembourg: Publications Office of the European Union, 2013.
